# Assessment of Knowledge and Attitude towards Blood Donation among Blood Donors in Jordan

**DOI:** 10.1101/2023.09.22.23295898

**Authors:** R.S. Omaish, Z.A. Al-Fayyadh, S.M. Al-Habashneh, S.Y. Al-Mashhdi, S.Y. Khasawneh, I.A. Naber, S.L. Bourghli, T.N. Al-Adily, A.M. Mahafzah, F.M. Al-Fararjeh, M.A Sughayer

**Author notes:** Corresponding author: Maher A. Sughayer MD.

## Abstract

**Objectives:** This study will identify blood donor characteristics in Jordan. The study aims to investigate the extent of awareness and knowledge regarding blood donation among blood donors and to explore their attitudes towards blood donation in general. It also highlights various motivational factors and obstacles which impact the decision to donate blood among Jordanian donors.

**Background:** Recruitment of low risk blood donors can be challenging. Efforts should be made to increase the level of awareness and positive attitude toward blood donation. An essential step to achieve this is obtaining comprehensive data about the current situation of awareness, knowledge, and attitudes of the population towards blood donation.

**Methods/Materials:** The present study was conducted at two blood donation centers in Amman, Jordan, during 2021. A total of 535 whole blood donors were included. Data regarding their demographic characteristics, blood donation history as well as their knowledge and attitudes regarding blood donation was collected by a questionnaire.

**Results:** Four hundred ninety participants (91.6%) were males while 45 only (8.4%) were females. One hundred forty subjects (19.4%) were first time donors while 431 subjects (80.6%) had previous donations. The participants’ median score in the knowledge section was 19.0 points (range 5-25 points). There was no significant correlation between participants’ overall attitude score and their corresponding demographic characteristics or their overall knowledge score.

**Conclusion:** Measures to improve knowledge and attitude toward blood donation should be implemented in order to meet the increasing demand for blood and blood components. Targeted campaigns, correction of some misconceptions, and using different motivations are suggested.

## Introduction

Blood is a major vital component of human body and safe blood transfusion is essential for improving health care and prevention of the spread of infectious diseases.

Despite countless years of studies and rigorous research, it does not seem to be promising that an ideal substitute for blood will be imminently found in the near future.^1^ Hence, the human blood donated is currently the sole option for replacement of blood and its various components.^2^

World Health Organization (WHO) estimated that 118.5 million blood donations are collected globally, 40% of these are collected in high come countries, home to 16% of the world’s population.^3^ It also estimates blood donation frequency to be 31.5 donations per 1000 people in high-income countries compared to 5.0 donations per 1000 people in low-income countries.^3^ The blood transfusion service in The Hashemite Kingdom of Jordan is done at 6 major sites. Donors in Jordan are either direct (e.g. relatives, friends and work colleagues) or voluntary non-remunerated donors.

Recruiting suitable donors for blood donations remains a challenge in both developed and developing countries.^4-6^ In view of the fact that voluntary unpaid donors comprise a much safer and a more reliable source of blood for transfusion compared to family replacement donors^3, 7^, efforts towards achieving self-sufficiency in terms of blood units must mainly focus on recruiting and retaining more volunteer donors in place of replacement donors. Several studies have been conducted to address awareness and attitude towards blood donation and suggested measures to enhance recruitment.^4-6, 8-13^ Analysis of 20 studies including 8,546 subjects concluded that being knowledgeable about blood donation was 2.85 times more likely to result into blood donation.^14^

Another study among health professions students including 598 students concluded that despite that 422 students (70.6%) didn’t take any courses regarding blood donation, 360 students (60.2%) had sufficient knowledge and 502 students (83.9%) showed high willingness to donate.^9^ A. Taş et al conducted a questionnaire based study on 220 medical students and concluded that the rates of blood donation and promoting people to donate blood were significantly higher in students who had received blood donation training compared to those who had not received such training.^15^

It should be noted that results obtained from different studies do not necessarily apply to our population due to different cultural, educational and socio-economic factors, along with varying perspectives across different populations.

Since reports on blood donation knowledge and attitudes in Jordan are lacking,^16^ the procurement of robust findings through execution of comprehensive studies is crucial before making any attempts towards growing population awareness and positive attitudes towards blood donation in the country.

In line with recent trends in blood donation research, this study aims to assess the knowledge and attitudes of Jordanian donors towards blood donation. It also proposes various motivational factors and obstacles impacting the process of blood donation in Jordan. Such information will form a solid base for the attainment of total voluntary blood donations, which are indeed the foundation of a sustainable blood supply.

## Materials and Methods

### Research design

The present research is based upon a cross-sectional, local population-based study which was conducted over a period of time extending from January through February 2021 at two blood donation centers in Amman, Jordan.

### Research approach and sampling method

The study includes 535 Jordanian participants. All participants were recruited from the blood banks directly affiliated to Jordan University Hospital (JUH) and King Hussein Cancer Center (KHCC), Amman, Jordan. Participants from both sites, exclusively involved whole-blood donors, were selected by convenient non-random sampling technique.

### Data collection method

All methods used in this study were approved by the Institutional Review Boards at the University of Jordan and King Hussein Cancer Center (KHCC), Jordan.

Prior to collecting data from participants at both sites, informed consent was obtained from the participants. It followed the guidelines set forth by the Institutional Review Boards of the University of Jordan and King Hussein Cancer Center, Amman, Jordan. Informed consent, written in the Arabic language, included a brief description of the study procedures, a discussion of the voluntary nature of the study, the right to withdraw without consequences, and the confidentiality of information.

Data was collected by means of an online survey sent out to participants at King Hussein Cancer Center. The survey needed an average of 13-15 minutes to be completed. A total of 111 responses were obtained from KHCC.

On the other hand, data was collected from participants at Jordan University Hospital by means of structured interviews conducted by the researchers. Researchers used the same online survey as the interview schedule. Interviews with the participants lasted for an average of 15-18 minutes. A total of 424 responses were obtained from JUH.

The survey questions were prepared following a review of earlier reports which probed the topics of knowledge and attitudes towards blood donation in various countries. Most survey questions required fixed-choice responses, though for some items there was the facility for brief text responses. The questions mainly covered 4 areas: participants’ demographic data, their blood donation history, their knowledge about blood donation, along with their attitudes and motivation towards blood donation.

### Data analysis method

Data from completed surveys was directly exported into a computer data sheet (Microsoft Excel).

The univariate analysis of the data was done using SPSS 25.0 for Windows. This yielded frequencies, corresponding percentages of the whole data, valid percentages, means, medians, standard deviations, as well as minimum and maximum values.

The bivariate analysis of the data was also done using SPSS version 25.0 for Windows. Knowledge was calculated based on 28 questions, in which a correct answer was given a score of 1, and any other answer was given a score of 0, including I do not know. The sum of the scores for each of the aforementioned questions yielded a knowledge total score out of 28 points.

Attitude was calculated based on 8 questions. Questions that had 4 options were given scores of 1, 2, 3, and 4. Questions which had 2 options were given scores of 1 or 4. A score of 4 was given to the answer with the most positive attitude while a score of 1 was given to the answer with the most negative. The sum of the scores for each of the 8 questions yielded an attitude total score out of 32 points.

Knowledge total score, attitude total score, and all the questions were correlated to ordinal and scale variables of the questionnaire using Spearman’s correlation. The level of significance was considered when p < 0.05. Knowledge total score and attitude total score were categorized to scores with a range of 5, and were then correlated to nominal variables using a Chi-squared test. The level of significance was considered when p < 0.05.

## Results

The study included 535 participants recruited from blood banks affiliated to Jordan University Hospital (JUH) and King Hussein Cancer Center (KHCC), Amman, Jordan. Out of the 535 study subjects, 490 (91.6%) were males while 45 only (8.4%) were females. About 75% of the population were younger than 40. In terms of education, 60.4% had a post-high school education. Additionally, 396 (74.0%) reported that they have previously done a blood test to know their blood type but only 375 participants (70.1%) said that their actual blood type matches the one stated on their national identity card.

One hundred and four subjects (19.4%) were first time donors while 431 subjects (80.6%) had previous donations (previous donors). Multiple previous donations (4 or more) were observed in more than 50% of previous donors. Among previous donors, voluntary donation was the most common motivation (66.1%) followed by direct donation for a family member or a friend (63.6%). The majority of previous donors have not reported to have had any complications post donation (81%).

Among first time donors, lack of awareness and/or having never been asked to donate before were the most common cause for not previously donating (51%). Table 1 summarizes the basic characteristics of participants and history of blood donations. Figures 1, 2 and 3 show the causes for previous donations, causes for not previously donating, and reported adverse effects post previous donations, respectively.

**Table 1:** Participants’ Demographic Characteristics and Their Blood Donation History (n=535)

| **Demographic Characteristics and Blood Donation History** |  | **Frequency (n)** | **Valid Percent (%)** |
| --- | --- | --- | --- |
| **Age (years)** | Less than 20 | 33 | 6.2 |
|  | 20-29 | 226 | 42.2 |
|  | 30-39 | 145 | 27.1 |
|  | 40-49 | 100 | 18.7 |
|  | 50-59 | 29 | 5.4 |
|  | Greater than 60 | 2 | 0.4 |
| **Gender** | Male | 490 | 91.6 |
|  | Female | 45 | 8.4 |
| **Marital Status** | Single | 269 | 50.3 |
|  | Married | 266 | 49.7 |
| **Level of education** | Below high school | 77 | 14.4 |
|  | High school | 135 | 25.2 |
|  | Diploma | 54 | 10.1 |
|  | University | 231 | 43.2 |
|  | Postgraduate | 38 | 7.1 |
| **Area of study (if applicable)** | Medical specialties | 41 | 13.4 |
|  | Scientific specialties | 130 | 42.3 |
|  | Literature and Humanitarian specialties | 129 | 42 |
|  | Arts | 7 | 2.3 |
| **Occupation** | Employed | 383 | 71.6 |
|  | Unemployed | 152 | 28.4 |
| **Monthly income (JDs)** | Less than 200 | 24 | 4.5 |
|  | 200-399 | 133 | 24.9 |
|  | 400-799 | 177 | 33.1 |
|  | 800-1199 | 49 | 9.2 |
|  | More than 1200 | 28 | 5.2 |
|  | No income | 124 | 23.2 |
| **Previous blood donation** | No | 104 | 19.4 |
|  | Yes | 431 | 80.6 |
|  | **Frequency of previous donations in previous donors** |  |  |
|  | Once | 81 | 18.8 |
|  | 2-3 times | 127 | 29.5 |
|  | 4-5 times | 67 | 15.5 |
|  | More than 5 times | 156 | 36.2 |

**Figure 1:**
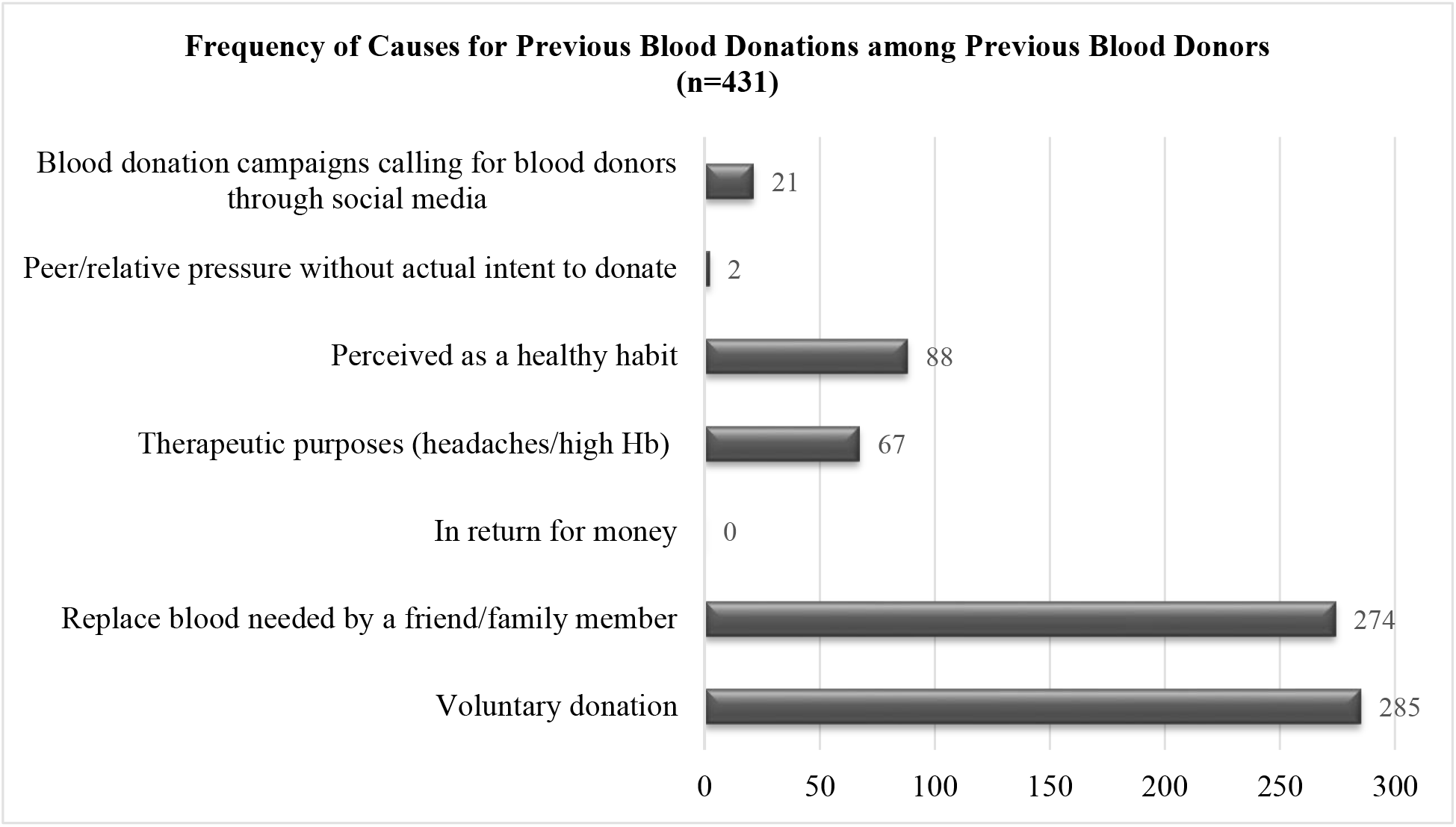
Frequency of Causes for Previous Blood Donations among Previous Blood Donors (n=431)

**Figure 2:**
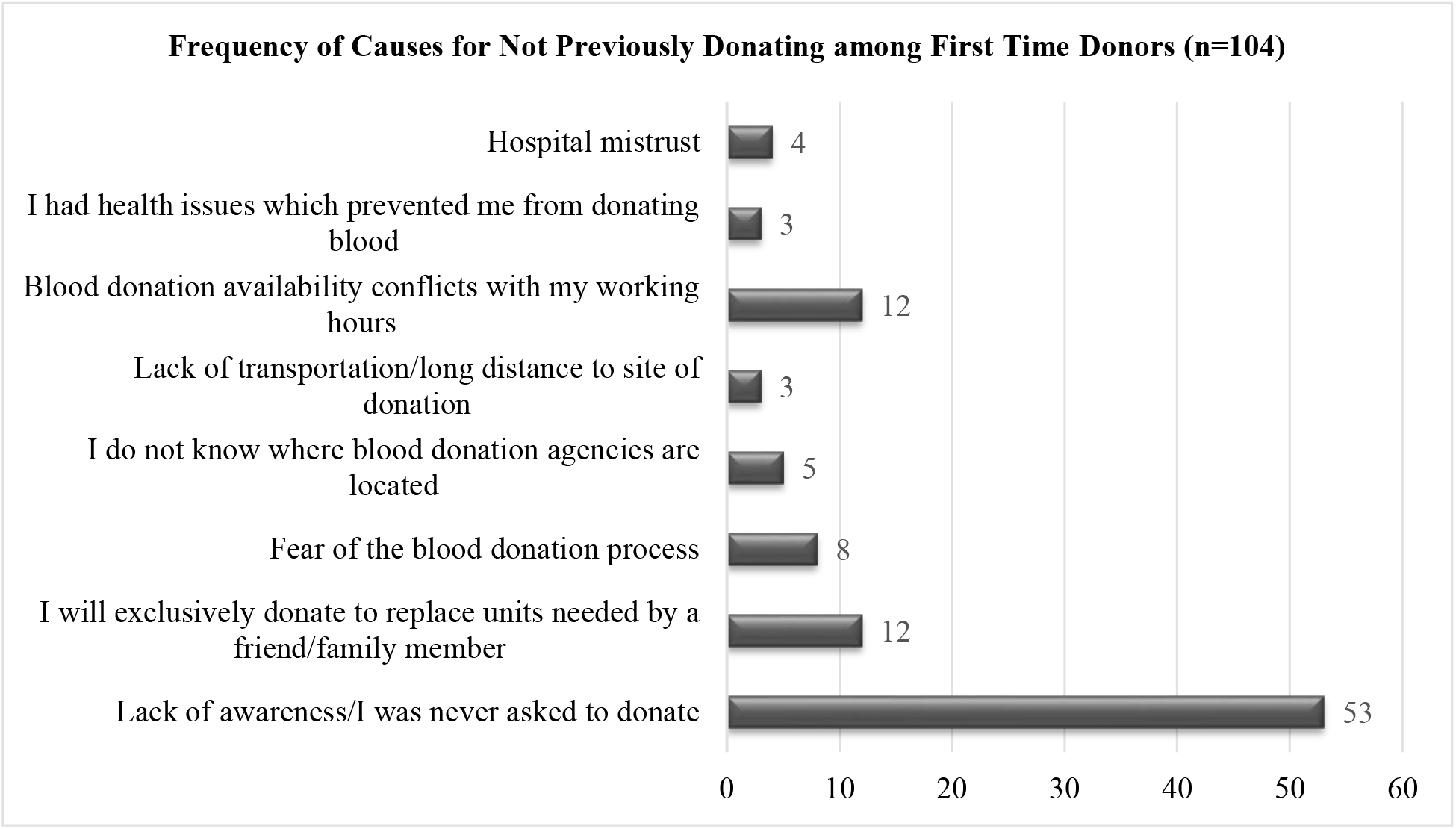
Frequency of Causes for Not Previously Donating among First Time Donors (n=104)

**Figure 3:**
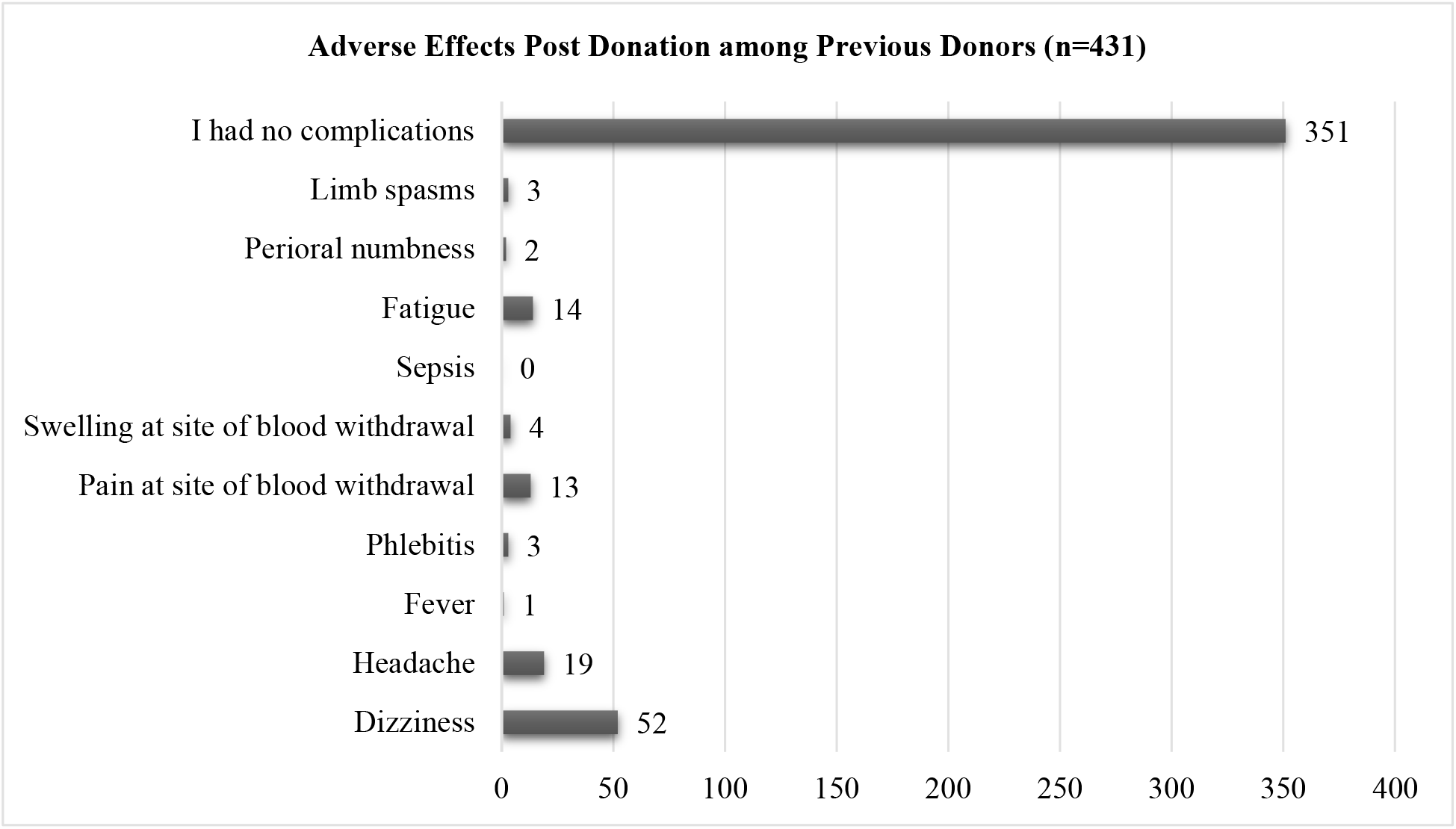
Adverse Effects Post Donation among Previous Donors (n=431)

### Knowledge

Knowledge was assessed based on a calculated result of 28 questions. The sum of the scores for each of the 28 questions yielded a knowledge total score of 28 points. The participants’ median score in the knowledge section was 19.0 points (range 5-25 points). Table 2 summarizes knowledge scores achieved by participants.

**Table 2:** Total Knowledge Scores.

| **Total Points** | **Frequency (n)** | **Percent (%)** |
| --- | --- | --- |
| **5-9** | 8 | 1.5 |
| **10-14** | 74 | 13.8 |
| **15-19** | 258 | 48.2 |
| **20-24** | 193 | 36.1 |
| **25-28** | 2 | 0.4 |
| **Total** | 535 | 100.0 |

More than half of the participants (53.1%) recognized that there are 4 major blood types and 75.9% knew that blood groups are genetically inherited. The majority of the participants (93.8%) thought that blood is screened before being given to patients, whereas only (68.6%) of them thought that blood gets processed (to separate different blood components) before being given to patients. Moreover, a significant proportion of the participants (85.6%) acknowledged that hepatitis B and/or C infection affects an individuals’ eligibility to donate blood. Upon asking whether patients can only receive blood from their first degree relatives, 484 participants (90.5%) disagreed.

Interestingly, 525 participants (98.1%) agreed that blood donation is safe for donors, and 515 participants (96.3%) agreed that blood donation has positive benefits upon donors’ health. Despite the fact that 521 participants (97.4%) agreed that blood units stored in blood banks are safe for use by patients, only 327 participants (61.1%) knew that blood units have an expiry date.

When it comes to blood banks status in Jordan in terms of blood unit sufficiency, 47.5% of the participants thought that blood banks in Jordan were short in terms of blood units, compared to 52.5% who thought blood banks had enough blood units or were not aware of the status, altogether.

Results of the bivariate analysis revealed that the overall knowledge score was not affected by participants’ age. However, knowledge was found to be significantly higher among female donors (P value <0.01).

In addition, participants with a higher level of education achieved a higher knowledge score (p value < 0.01). Interestingly, donors with a higher level of education recognized more than others that blood units stored in blood banks have an expiry date and that blood banks in Jordan lack sufficient blood units (both with a p value of < 0.01). Also, participants who have achieved a higher level of education were more likely to have previously donated blood because they thought it was healthy and in response to blood donation campaigns calling for blood donors through social media (both with a p value of < 0.01).

In regards to occupation, employed participants were found to be more knowledgeable about blood donation in general (p value < 0.01).

More interestingly, participants who have ever participated in voluntary activities in their lifetime were found to achieve higher scores in the knowledge questions (p value < 0.01).

Finally, our questionnaire did not detect a statistically significant difference between first time and previous donors when it comes to the total knowledge score. However, first time donors, who have not donated blood previously because they claimed that they would only do so to replace blood units needed by family members or friends, were more likely to think that blood donors and recipients should be first degree relatives (p value < 0.01).

### Attitude

Attitude was assessed based on a calculated result of 8 questions. The sum of the scores for each of the 8 questions yielded an attitude total score of 32 points. The participants’ median score in the attitude section was 25.0 points (range 12-32 points).

Participants’ views on blood donation in terms of it being an ethical act, a religious or national duty and a healthy habit are summarized in Table 3.

**Table 3:** Participants’ views on blood donation.

| **View on**<br>**blood**<br>**donation** | **Participants' response (%)** |  |  |  | **Percent (%)** |
| --- | --- | --- | --- | --- | --- |
|  | **I strongly**<br>**agree** | **I agree** | **Neutral** | **I disagree** |  |
| **Ethical act** | 72.9 | 25.0 | 1.5 | 0.6 | 100.0 |
| **Religious**<br>**duty** | 49.5 | 34.0 | 11.8 | 4.7 | 100.0 |
| **National**<br>**duty** | 54.0 | 34.8 | 7.1 | 4.1 | 100.0 |
| **Healthy**<br>**habit** | 53.2 | 38.9 | 5.8 | 3.0 | 100.0 |

When it comes to motivational factors, a great percentage of the participants (77.8%) agreed that mobile blood donation caravans roaming around public areas will further motivate them to donate blood while almost two thirds agreed that both getting free blood tests (67.5%) in return for blood donation or getting a day off work (67.3%) will make them more likely to donate blood in the future. On the other hand, about 9.5% agreed that receiving monetary compensation in return for blood donation will increase their likelihood of donating blood.

Results of the bivariate analysis revealed there was no significant correlation between participants’ overall attitude score and their corresponding demographic characteristics (including age, gender, level of education, area of study, occupation and monthly income) or their overall knowledge score.

However, older participants were more likely to agree that blood donation is both a national duty and a religious duty (both with a p value < 0.01). In addition, participants who achieved higher levels of education were more likely to agree that blood donation is both an ethical act and a healthy habit (both with a p value < 0.01). Interestingly, participants with higher income were more likely to agree that blood donation is an ethical act, a religious duty, a national duty and a healthy habit (p value < 0.01).

## Discussion

Our study showed that the majority of blood donors are males. This is consistent with previous reports from the Kingdom of Saudi Arabia and Nigeria.^10, 17^ On the other hand, reports from Iceland and Germany showed a more even distribution.^18, 19^ Despite the fact that female participants only comprised 8.4% of our study population, they were found to be more knowledgeable of blood donation in general when compared to male participants. This can be attributed to the fact that medical-related professions were more common among female donors when compared to male donors (35.6% and 5.5%, respectively).

Only 24.5% of our study population were older than 40 years of age. Older first time donors were more likely to have not donated blood previously due to thoughts of having health issues affecting their eligibility to donate. This highlights the need for a better understanding of the possible interfering medical conditions and better assessment of older adults for fitness for blood donation.

The rate of first time donors among different populations is variable. Suemnig A et al reported a first time donor percent of 14.3%, while Niazkar HR et al reported a percent of 26.5%.^20, 21^ In addition, a previous study conducted among the Jordanian population revealed a first time donor proportion of 25.4% of the total study population.^16^ In our present study, about one fifth (19.4%) of the participants were first time donors, while the remainder (80.6%) were previous donors.

In our study, more than half of previous donors had 4 or more previous donations. The most common cause behind previous donations found among our participants was voluntary donation. This is consistent with previous reports which found altruism and the wish to help others to be the most common driving force behind blood donation.^13, 22^

It has been reported that proper knowledge of blood donation was indeed an important factor for donating blood. In a study conducted among the Saudi population, Alfouzan N reported that more knowledgeable subjects tended to donate blood more than those of lower levels of knowledge.^13^ Interestingly, first time blood donors from our study population who reported not having donated blood before because they would only do so to replace blood units exclusively needed by their family members were more likely found to believe that blood donors and recipients should be first degree relatives. This indicates that correction of such a misconception may alter donation attitudes among the Jordanian population.

In contrast to studies conducted in Tanzania^23^ and Nigeria^24^ which inferred that voluntary donations were correlated to secondary school education, our study did not detect a correlation between participants’ attitude towards blood donation and their corresponding level of education. However, in our study, participants with a higher level of education were found to be more knowledgeable of blood donation in general, as they were found to achieve relatively higher total knowledge scores than those with lower levels of education.

A strong ethical and religious drive was noted among our study population. In accordance with previous studies among the Saudi population,^10, 13^ most participants from our study agreed that blood donation is an ethical act, and both, a religious and a national duty. In fact, in our study, this drive was mainly observed among older participants, as well as those with a higher level of education and a higher income.

A previous report on barriers to blood donation in Jordan^16^ revealed that participants having not received blood when needed was the single major barrier affecting blood donation among the Jordanian population, comprising 78.4% of the participants’ answers. This was followed by side effects of blood extraction (18.8%), having health problems (9.6%) and fear from blood (7.8%). In our present study, lack of awareness regarding blood donation and/or having never been asked to donate blood before were the predominant barriers (50.9%) to previous blood donation among first time donors, followed by direct donation to exclusively replace blood units when needed by family members or friends (11.5%), blood donation conflicting with participants’ working hours (11.5%) and fear of the blood donation process in general (7.7%).

It has been previously reported by Thomson et al that 80% of first time donors globally would never return to donate.^25^ Therefore, the efficacy of various methods used in attempts to increase the return rate of first time donors was assessed in previous studies. Hashemi S et al found that different interventions, including phone reminders, educational and emotional letters, along with motivational meetings, were effective in improving the return rate.^26^ Among our study population, donors thought that their likelihood of donating blood in the future would increase if mobile blood donation caravans were available in public areas, and if they get free blood tests or a day off work in return for their donation.

On the other hand, monetary compensation was disfavored as an incentive to blood donation in several previous studies. A study which probed the factors motivating the Nigerian population towards blood donation found that only 6.46% of the respondents would donate blood in return for money.^24^ Similarly, a study conducted in the United States of America to assess blood donor attitudes towards various incentives reported that 7.3% of the donors would be discouraged from donating if they received cash.^27^ Data obtained from our study was much in line, as 90.5% of our participants objected donating blood in return for money.

In the Hashemite Kingdom of Jordan, citizens’ national identity (national ID) card includes 18 data fields, one of which is the holder’s blood type. In our study, only 396 participants (74.0%) reported that they have ever done a blood test to know their blood type. A close percentage of the participants (70.1%) reported that their actual blood type matches the one stated on their national identification card. This discrepancy in reporting calls for the need of applying more strict measures to ensure the correct documentation of blood groups on national ID cards. Indeed, knowledge of the frequency distribution of the ABO blood groups is essential for blood bank information as well as safe transfusion of blood in cases of emergency.^28^

## Conclusion and Suggestions

Blood donation is an essential component of patients care and efforts should be made to increase recruitment of voluntary donors and increase return rate of first donors. Based on our findings among blood donors at JUH and KHCC, we suggest:

- Blood donation awareness campaigns need to target people with least knowledge toward blood donation found in our study, such as the unemployed and those who have not completed their education beyond high school. Programs such as school educational courses or school visits to blood banks are suggested.
- Females comprised only 8.4% of total donors’ population. Measures to encourage females to participate should be implemented. Television advertisements or campaigns to target university students or health club members can be of help.
- Correction of misconception or inaccurate information such as that blood units have no expiry date or that blood donors and recipients should be first degree relatives. More than half of donors thought that blood banks in Jordan had enough blood units or were not aware of their status. Efforts should be made to change this inaccurate information.
- Consider designing and starting a blood donation motivation scheme. Motivations such caravans, free blood tests and days off work were thought by our study population to increase their likelihood of donation.

## Data Availability

All data produced in the present study are available upon reasonable request to the authors

## Acknowledgements

The authors thank the staff at both blood donation centers.

